# How engagement with an online health community can influence self-management behavior: insights from a qualitative analysis of a UK asthma online community using Behavior Change Techniques taxonomy

**DOI:** 10.1101/2025.08.11.25333420

**Authors:** V Dhir, HE Wood, X Li, GD Karampatakis, CJ Griffiths, A De Simoni

## Abstract

**Background:** Patients with long-term conditions take part in online health communities (OHCs) for advice and guidance from peers. Engagement with OHCs may prompt changes in self-management behavior. Limited information exists on determinants of behavioral change resulting from engagement with an OHC.

**Aims:** Firstly, to identify whether engagement with an OHC results in behaviour change. Secondly, to identify the behavior change outcomes, and whether specific behavior change techniques were used.

**Design & Setting:** Qualitative analysis of posts written between December 2022 and August 2023 in the asthma OHC of the Asthma + Lung UK (ALUK) charity.

**Method:** The search term ‘helped’ was used in the Google search engine to highlight relevant posts and threads. Two researchers read through these posts to identify threads that described behavior change as a result of OHC engagement and classified them based on the specific actions taken or intentions expressed by users. Earlier posts in the same threads that encouraged behavior change were classified using Michie’s Behavior Change Technique (BCT) Taxonomy to identify potential contributing techniques.

**Results:** Thirty-five threads were identified, with 703 posts contributed from 200 users. Users’ characteristics were mostly hidden. Seventeen (48.6%) of these threads included posts where the users who started the thread went back to it to describe their behavior change as a result of engaging with the OHC. Within these threads, 6 posts demonstrated actual behavior change, 32 posts intention to change behavior, and one post both intention and actual behavior change. Behavior change outcomes included: contacting primary care services; trying out other users’ recommendations; consulting a healthcare professional to change medication; and purchasing equipment.

Fifty-two distinct posts (out of a total of 362 posts) by 32 users were identified as prompting behavior changes, through the use of 12 out of 96 available BCTs. Common BCTs employed by users were: ‘Demonstration of behavior’ (n=30), ‘Action planning’ (n = 26), ‘Instruction on how to perform the behavior’ (n=17), ‘Adding objects to the environment’ (n=15), and ‘Pharmacological support’ (n= 10). All threads where a behavior change was prompted, contained 2 or more BCTs. Eighteen of the 200 users were very active (i.e., superusers), contributing at least 10 posts during the 9-month period of the study. 9/18 superusers were among the 32 users who posted in threads where actual behavior change or intention to change behavior was observed.

**Conclusion:** Engagement with an OHC can influence the self-management behavior of patients with long-term conditions. Recognized BCTs underpinned threads demonstrating self-management behavior change.

## Introduction

Asthma is a highly prevalent chronic lung disease: in 2019 it affected 262 million people worldwide and caused 455,000 deaths [1]. In 2018, 17% of men, and 18% of women were the proportion of adults with diagnosed asthma in the UK [2]. While asthma cannot be cured, it can be managed effectively with medication, education and acquisition of self-management skills [2,3].

The British Thoracic Society (BTS), Scottish Intercollegiate Guidelines Network (SIGN), and National Institute for Health and Care Excellence (NICE) joint guidelines ‘Asthma: diagnosis, monitoring and chronic asthma management’ suggests offering self-management interventions to people with a diagnosis of asthma [4]. One of these interventions may include IT based monitoring and education [4]. The acquisition of self-management skills by patients with asthma can reduce emergency use of healthcare resources, including emergency department (ED) visits, hospital admissions and unscheduled primary care consultations [5]. Ability to self-manage can also improve markers of asthma control, reduce symptoms and work absence and improve quality of life [5].

In a survey of 3001 adults in the US, it was found that approximately 18% of internet users have gone online to find others who might have health concerns similar to theirs [6]. Patients with long-term conditions can access information on their health needs from other users with similar health conditions through online health communities (OHCs). Online social network-based health interventions can either use standalone social networks such as the Asthma + Lung UK (ALUK) charity asthma community forum [7], or have interventions developed for more popular social media platforms [8] such as *Facebook* [9] or *X* (formerly *Twitter*) [10]. The ALUK asthma OHC has currently over 21,000 registered members and 24,000 posts (as of May 2024) and is used by patients with asthma and their carers [7]. OHCs, such as ALUK, allow users to read and reply to posts, interact with other users and access resources quickly [11–13]. OHCs have the potential to positively change patient behavior by offering a supportive environment in which information and advice are freely exchanged amongst peers (other patients, and/or carers of patients, with similar conditions) [14–15].

Appropriate behavior change is a critical component of effective self-management for patients with long-term conditions, as it empowers individuals to take an active role in managing their health through lifestyle modifications, adherence to treatment, and symptom monitoring. Sustainable behavior change in the context of self-management often requires addressing various psychological, social, and environmental factors that influence patients’ daily decisions and habits. *Behavior change interventions that are strongly based in theory have been found to have a greater impact than those that were not [16].* These interventions that redirect or alter behaviour can be complex to record, and therefore difficult to use in research [17]. The Behavior Change Technique (BCT) Taxonomy developed by Michie et al.[17] provides a comprehensive framework for systematically studying and promoting behaviour change. It contains 93 distinct, and consensually agreed BCTs which are used to specify interventions [17], allowing the BCTs to be observable and replicable [17]. These BCTs can be single, or used in combinations [17]. By recording BCTs used in threads where a behavioural change has occurred, we will identify frequent BCTs, and their combinations which may lead to behavioral change as a result of engagement with this online health community.

Most existing literature on behavioral change as a result of engagement with OHCs focuses on health behaviors such as smoking cessation or increasing physical activity, rather than self-management of chronic conditions [18–20]. The aim of this study is to investigate whether engagement with an asthma OHC led to any behavior change and to determine which BCTs, if any, were employed by users to facilitate this change. By identifying the specific techniques used, this study seeks to provide a deeper understanding of the nature of self-management behavior change as a result of OHC engagement.

## Methods

We conducted a qualitative analysis of posts written publicly in the ALUK asthma OHC using the BCT taxonomy. The ALUK OHC was chosen due to our previous work showing a wealth of experience being exchanged by users on this forum [21,22]. Box 1 clarifies terms used in the current study.

### Box 1. Definition of OHC terms

|  |  |
| --- | --- |
| User | An individual who has signed up with the OHC. |
| Username | Moniker adopted by the user to identify themselves. |
| Post | A comment/reply/question etc ‘posted’ on the forum by a user. This can be to start a new ‘thread’ on a particular topic, or in reply to or commenting on a previous post. |
| Thread | A group of posts about a particular topic, with a title, often started by a user posting a question or comment, followed by replies from other users. |

### Ethical issues

Ethical aspects of this study were assessed by Queen Mary University of London’s Ethics of Research Committee. The study was exempt from full review (QMERC22.279). The passive analysis approach used in this study is generally considered non-intrusive [23]. To protect the identity and intellectual property of OHC users, summative descriptions of quotes were used. Permission for data collection and analysis was sought and granted by both HealthUnlocked (our industry partner, which hosts the OHC platform) and ALUK before starting the study.

### Data Collection

To provide a manageable amount of data for analysis, data extraction was limited to content posted between 1 December 2022 and 31 July 2023. Google search engine was used to look for relevant posts within the Asthma UK community platform. Different search keywords were tested to determine which would be most useful in obtaining relevant posts on the OHC. The keyword ‘helped’ was found to retrieve threads describing behavioral change. All posts within the relevant threads were chronologically extracted and exported into MS Excel to create the dataset for analysis. Excel was used to organize data and calculate descriptive statistical measures (mean, SD).

### Data Analysis

*Two researchers (VD and HEW) independently identified posts that described behavior change, highlighting actual changes in behavior as well as an intention to change behavior. VD is a general practitioner working regularly with asthmatic patients. HEW is an academic researcher with experience and training in qualitative analysis techniques. VD and HEW independently read through all the posts, and classified those that prompted behavior change using the BCT Taxonomy [17].* The researchers then met to resolve any disagreements. Each post was assigned the appropriate BCT codes from the 93 possible hierarchically-clustered techniques. Posts describing intention or actual behavior change were then classified in terms of behavioral outcomes: e.g., contacting a health professional, following self-supportive advice, etc. Posts were also reviewed to see whether the user starting the thread was the same user who demonstrated the behavior change, or whether other users within the threads also demonstrated a behavior change.

Data were analyzed to understand how active the users were who contributed within these threads.

Posts were also read to ensure appropriateness of advice given, and where applicable, the moderation of these posts by forum moderators. Posts were read for advice that could be considered controversial, potentially harmful or contradicted the BTS/SIGN/NICE joint guidelines [4].

## Results

### Overview

Using the search term ‘helped’, we identified 35 threads, including a total of 703 posts contributed by 200 individual users. Out of the 703 total posts, three were from users who had chosen to hide their usernames, making them unidentifiable. Threads were initiated by 33 unique users.

Among the 200 identified users, the majority (n=158/200, 79%) contributed fewer than five posts each, while a small number of more active users (n=18/200, 9%) contributed 10 or more posts. Notably, the most prolific contributor was a member of the forum moderation team, labelled as ‘Administrator’. Figure 1 ‘Consort diagram’summarizes the data used in this analysis.

**Figure 1.**
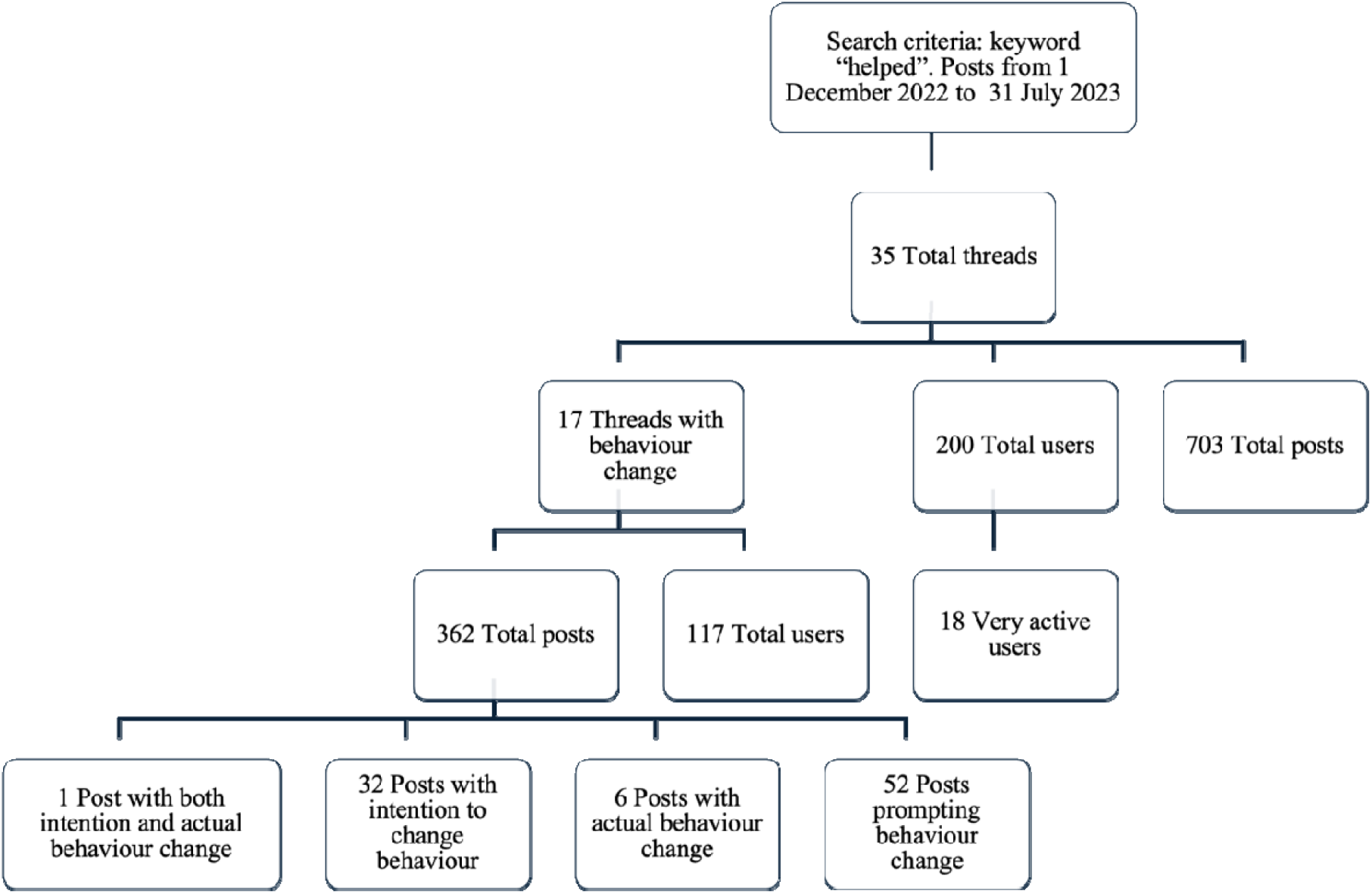
Consort diagram: Data retrieval

### Threads including behavior change

VD read through all 35 threads containing the keyword ‘helped’ and identified 30 posts in which the user demonstrated an intention to change and 7 demonstrated actual change. HEW also read through all threads independently, and identified 32 posts with intentions and 7 with actual changes. These differences were subsequently discussed and resolved following a meeting.

It was agreed that out of the 35 threads, 17 (comprising a total of 362 posts by 117 individuals) included at least one post in which the original user returned to update on their progress, describing a change in behavior or an intention to change.

Overall, 18 users (18/117, 15%) expressed an intention to change behavior or an actual behavior change in 39/362 posts (11%). In two threads, a post from a user other than the thread starter indicated an intention to change behavior. The majority of these posts (n=32/39, 82%, 15 threads) described intention to change behavior, while six (6/39, 15%, 5 threads) an actual behavior change. Only one post (3%) included both an intention and an actual change in behavior. Table 1 gives details of the 17 threads in this subset.

**Table 1.** Details of the 17 threads that included at least one post indicating a behavior change or intention to change behavior, from a total of 35 threads including the search term ‘helped’.

| **Subject of thread** | **User indicates intention to change** | **User indicates actual change** | **Number of posts in thread<sup>a</sup>** | **Number of posts prompting change** | **BCT used** |
| --- | --- | --- | --- | --- | --- |
| Silent reflux and asthma | x |  | 19 | 1 | 4.1, 5.1, 6.1 |
| Asthma and COVID | xx<sup>b</sup> | x | 28 | 3 | 1.4, 5.1, 6.1, 6.2, 11.1 |
| Sore throat and inhalers | x |  | 23 | 5 | 1.4, 4.1, 5.1, 6.1, 8.1, 9.1, 11.1 |
| Not being able to exhale fully |  | x | 10 | 4 | 1.4, 5.1, 6.1, 11.1 |
| Burning sensation in lungs | x |  | 8 | 2 | 1.4, 5.1, 6.1 |
| Using Fostair and Ventolin | x |  | 15 | 1 | 1.4, 4.1 |
| Dog dander | x | x | 15 | 1 | 4.1, 6.1, 11.1 |
| Salamol vs. Ventolin | x | x | 32 | 8 | 1.4, 2.7, 4.1, 6.1, 9.1 |
| Tezepelumab/ Spirometry with Reversibility | x |  | 15 | 2 | 1.4, 4.1, 6.1 |
| Chronic infections | x |  | 23 | 1 | 1.4, 6.1 |
| Air purifiers | xx<sup>b</sup> | x | 34 | 13 | 4.1, 6.1, 12.5 |
| Side effects of Clenil Modulite | x |  | 28 | 3 | 1.4, 6.1 |
| Child with asthma | x |  | 12 | 2 | 1.4, 3.2, 4.1 |
| Powder inhalers | x |  | 37 | 2 | 1.4, 3.1, 6.1 |
| Mannitol challenge | x |  | 36 | 1 | 1.4, 4.1, 6.1 |
| Oral steroids | x |  | 13 | 1 | 1.4, 6.1, 11.1 |
| Fostair use | x | x | 16 | 2 | 1.4, 3.1, 9.1, 11.1 |

<sup>a</sup> Including original post starting the thread; <sup>b</sup> not the user who started the thread; BCT, behavioral change technique

Out of 39 posts indicating behavior change or intention to, 22/39 (56%) had sought or planned to seek help from a healthcare professional, including via contacting the ALUK helpline (which is staffed by specialist respiratory nurses), 16/39 (41%) had used self-management behaviors or planned to, and 3/39 (8%) stated they intended to try self-management first, before contacting a HCP if this approach wasn’t effective. Self-management behaviors included users planning to be more forceful in their communications (2/16, 13%), trying a recommended method of self-care/self-monitoring of symptoms, an over the counter (OTC) medication or non-pharmaceutical remedy, or a change to their environment to avoid asthma triggers (11/16, 69%), and planning to do research (3/16, 19%). Specific examples of posts describing each of these types of behavioral changes are given in Table 2 using paraphrased quotes.

**Table 2.**
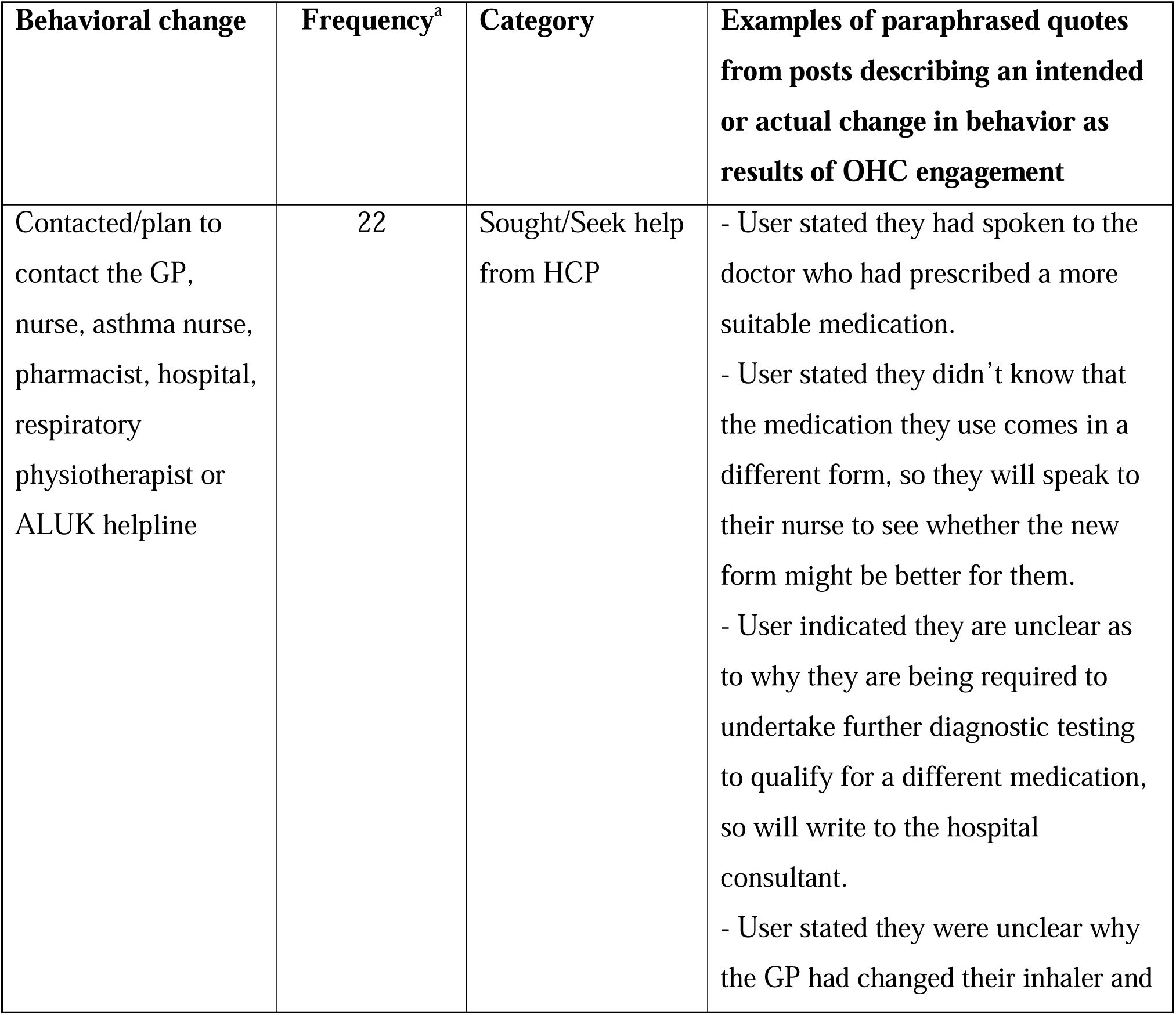
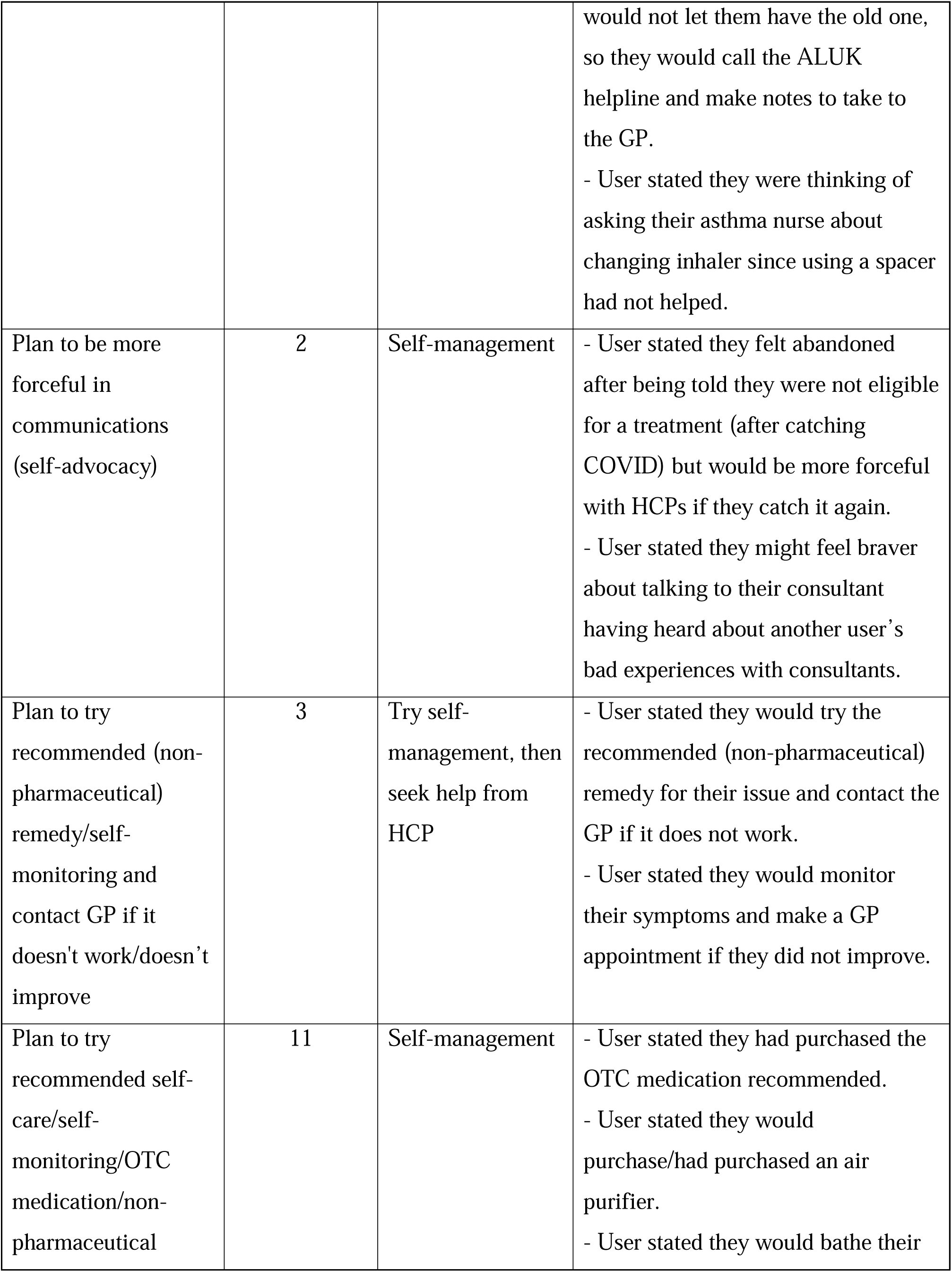
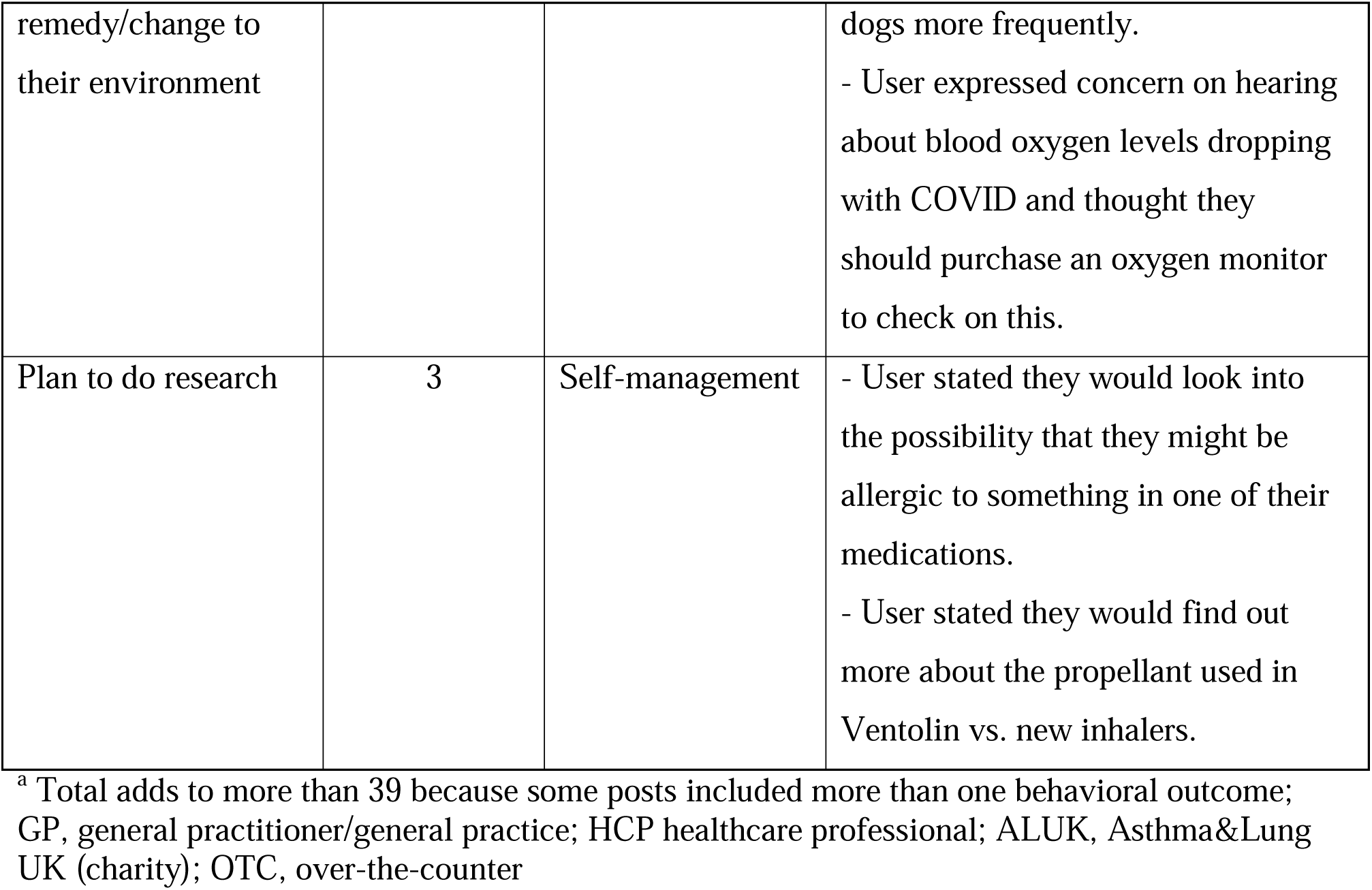
Types of actual or intended change in behavior resulting from engagement with the asthma OHC.

### BCTs triggering behavior change

We identified 52 posts deemed instrumental in prompting behavioral change or intention to change behavior, which were contributed by 32 individual users (see Table 1). There was very little overlap between users prompting behavior change and those reporting an intention or actual change in behavior, with only two users both indicating an intention to change their own behavior and prompting behavior change in another user. The 52 posts prompting behavior change were classified using the BCT taxonomy [17].

A total of 12 different BCTs were identified within these posts (see Table 3), with the most frequently observed techniques being ‘Demonstration of the behavior’ (used in 30/52 posts, 58%) and ‘Action planning’ (used in 26/52 posts, 50%). Table 3 gives examples of posts employing each of the 12 different BCTs identified, using paraphrased quotes to protect the identity of users.

**Table 3.**
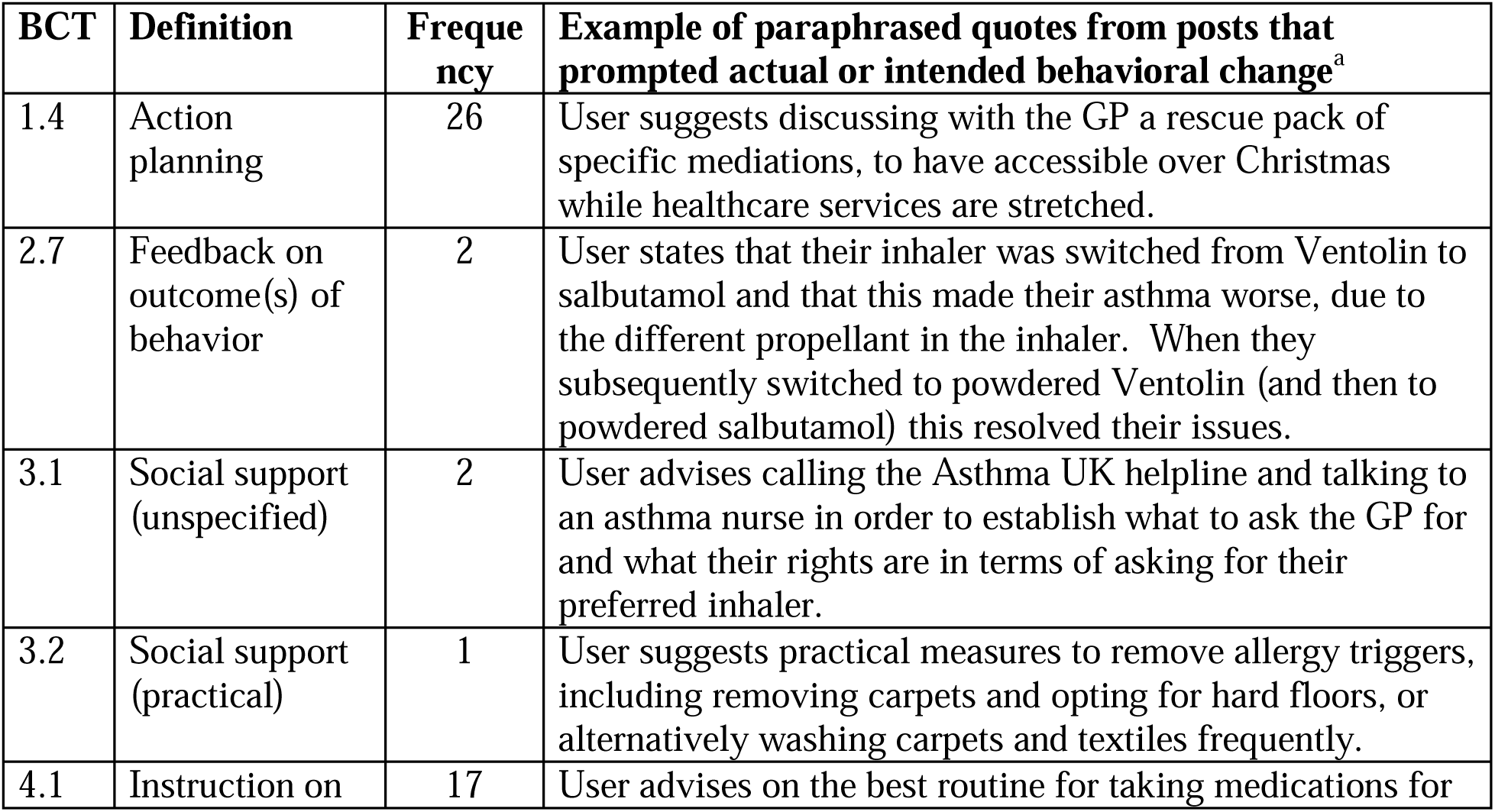
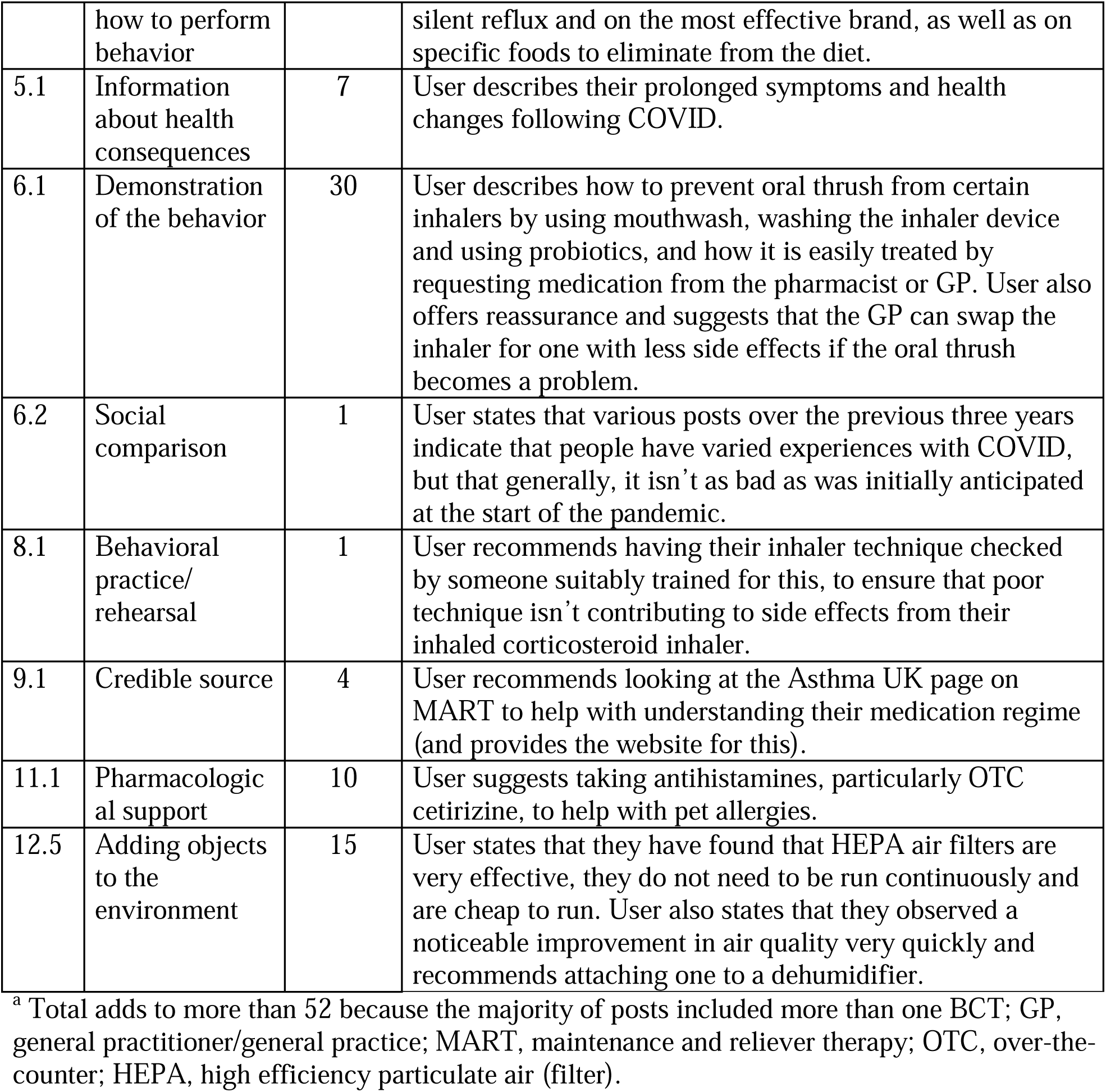
Behavior change techniques (BCTs) used in posts identified as prompting actual or intended behavior change (from a total of 52 posts)

### Appropriateness of advice

None of the identified posts which prompted actual/intended behavior change gave advice/recommendations that could be considered controversial, potentially harmful, or contradicted the BTS/SIGN/NICE joint guidelines [4], indicating that the advice shared on the asthma OHC is generally safe. However, within the 17 selected threads there were three posts which mentioned or recommended the use of *Olbas oil* [24], which is not included in the BTS guidelines. No users replied to these posts, apart from a member of the forum moderation team who replied to one, advising users against the use of *Olbas oil*, pointing out that it can be a trigger for asthma [25].

## Discussion

### Summary of findings

This study provides qualitative evidence that engagement with an asthma OHC drives self-management behavioral change as well as the intention to change behavior. In addition, users demonstrated a range of behavior change outcomes. These behavior changes were prompted as a result of discussions with other members of the OHC. The posts prompting the behavioral changes were categorized using BCTs. We observed that threads that resulted in behavior change contained multiple BCTs.

The most common behavior change demonstrated or intended was to seek help from a healthcare professional (HCP), including calling the ALUK charity helpline. Determinants of self-management behavioral change were characterized, encompassing 12/93 BCTs. The most common BCTs to trigger self-management behavior change were ‘Demonstration of the behavior’ and ‘Action planning’.

These findings suggest that patients may not always realize when they need to seek medical advice, but engagement with an OHC helped them to recognize when seeking medical advice was appropriate, versus managing their own symptoms. Examples of situations in which users appeared unsure of whether or not to seek medical advice included: being unsure about their symptoms and how serious they were; whether prescribed medication was appropriate; experiencing side effects of medication; COVID-19 infection/symptoms; receiving confusing advice from different HCPs; and regarding different types of asthma medications. The posts which prompted users to change their behavior included directing users to contact a doctor, asthma nurse, pharmacist, hospital consultant, respiratory physiotherapist, or the ALUK helpline, mainly around issues about their medication.

On the other hand, advice and recommendations from peers also helped users to follow self-management methods, rather than contacting a HCP. Examples of recommended self-management methods included: taking an OTC remedy or trying a non-pharmaceutical remedy; purchasing equipment or making other changes to their home environment; self-monitoring symptoms; and undertaking their own research into a particular issue. These findings suggest that the peer support available within this OHC may be beneficial in supporting self-management of asthma. This could help to reduce the strain on healthcare services by educating users about alternatives to contacting a HCP and encouraging them to try these self-management methods instead of or before seeking professional help, only doing so when absolutely necessary.

We did however consider the safety aspect of the information being shared in the OHC. We found active and frequent participation by moderators to ensure safe guidance was being issued. From a study undertaken by Wood et al [26], examining the role of the moderators of the ALUK asthma forum, we are aware that new posts are monitored daily, and any posts not adhering to the forum’s guidelines are quickly dealt with e.g. a moderator editing the post to add advice or clarification, or taking down the post and any replies to it. However, we are aware that prior to posts being moderated, it is possible for a user to post advice on the forum that could potentially cause harm. For example, if a user was recommended a natural remedy which they were allergic to. Therefore, online discussion boards should still be approached with caution.

We also recognize it is possible that OHCs such as the ALUK asthma OHC may attract more motivated individuals to join the forum, and be more willing to change behavior compared to an average person living with asthma.

One systematic review states that standalone OHCs can be helpful to the users they retain over time. However, retention of users of OHCs is generally poor compared to social media platforms [8] such as *Facebook* [9]. *Facebook* is an example of how people most commonly use social media platforms. Users of *Facebook* commonly interact with other users they may already have an offline connection to, whereas users of standalone OHCs are often strangers to each other [8]. Behavioral change interventions involving users who already know each other may increase engagement [8].

Overall, this study shows that engagement with this OHC promotes self-management behaviors and supports users to recognize when to seek medical advice. Behavior changes were prompted by peer interactions which contained BCTs. The study demonstrates that this OHC provides valuable support to people with asthma, however, we acknowledge the effectiveness of OHCs may vary depending on factors such as moderation, user retention, and motivation.

### Strengths and limitations

A strength of this study is that it is based on real-world data, focusing on behavioral change within the context of an asthma OHC. However, the generalizability of these results to other OHCs for asthma or other long-term conditions may be limited. The ALUK asthma OHC is a long-standing forum and is moderated by ALUK specialist nurses and patient moderators.

Other online communities for patients with asthma or other long-term conditions may not be moderated in the same way and therefore may not lead to positive behavioral changes. Moreover, determinants of self-management behavioral change associated with different medical conditions may well differ.

Another strength of this study is the use of BCT taxonomy, which adds a structured approach to understanding the techniques employed within the asthma community to influence behavioral change. However, qualitative analysis is subjective and different researchers may identify different BCTs. During the process of applying BCTs to the posts, we encountered the challenge of converting subjective, and unstructured data, into their appropriate categories. This process of categorizing, reduces subjectivity. However, in our analysis, we found close agreement between researchers in the BCTs identified. It is also worth considering that while we have identified BCTs within specific threads of discussion, these techniques may not comprehensively capture the complex interplay of factors influencing users’ decision making, and consequently behavioral change.

Another limitation of conducting research using content posted on OHCs is that it relies on self-report by users, which may be subject to bias such as social desirability or misrepresentation of experiences. As each user’s perception of their illness is unique, overgeneralization can negatively impact treatment pathways and the effectiveness of relief, highlighting the need for a balanced approach that preserves individuality while organizing data,

The search term ‘helped’ could be a further limitation as it may exclude search results that may have been found if other similar terms may have been used (examples include ‘improvement’). However, we found a large amount of relevant data returned using the search term ‘helped’, and this data had to be further filtered by date range to make it more manageable. We may also have omitted from our analysis users who read and commented on older posts or indeed passive users who may have changed or intended to change their behavior as a result of engaging with the asthma OHC, but did not reply or post on the forum to evidence that change or intention. Information on participant age, length of time with asthma, severity of asthma, and demographics would have provided further insights into the type of users participating in behaviour changes on this OHC. However, this data was not publicly available.

### Comparison with existing literature

Theoretical models of behavior change help improve traditional health promotion as well as promotion involving social media [16]. Interventions incorporating a greater number of BCTs, such as those observed in this study, tend to have larger effects than those studies with fewer BCTs [16]. It is hypothesised that different techniques address different aspects of the behaviour change process [16], and this may help explain the use of multiple BCTs leading to behavioral change in this study.

The BCTs can be mapped onto Dennis’s [27] conceptual model, a theoretical framework for developing, measuring, and evaluating healthcare interventions. This model, adapted from Social Support Theory [28], has been tailored by Dennis to the healthcare context and specifically to peer support interventions.

Dennis’s conceptual model [27] has been used as a logic model (Figure 2 ‘logic model’) to guide the development of intervention and formulate hypothesis regarding potential improvements in patient long-term outcomes [29]. The framework describes the key attributes of peer support being emotional, informational and appraisal. It also specifies the mechanisms through which peer support is hypothesised to produce beneficial outcomes, alongside required antecedents for effective implementation, such as appropriate moderator training (see figure 2 ‘logic model’). This theoretical framework was selected because it articulates the mechanisms through which peer support interventions influence health outcomes within the broader programme of research of which this work is a part [29,30].

**Figure 2.**
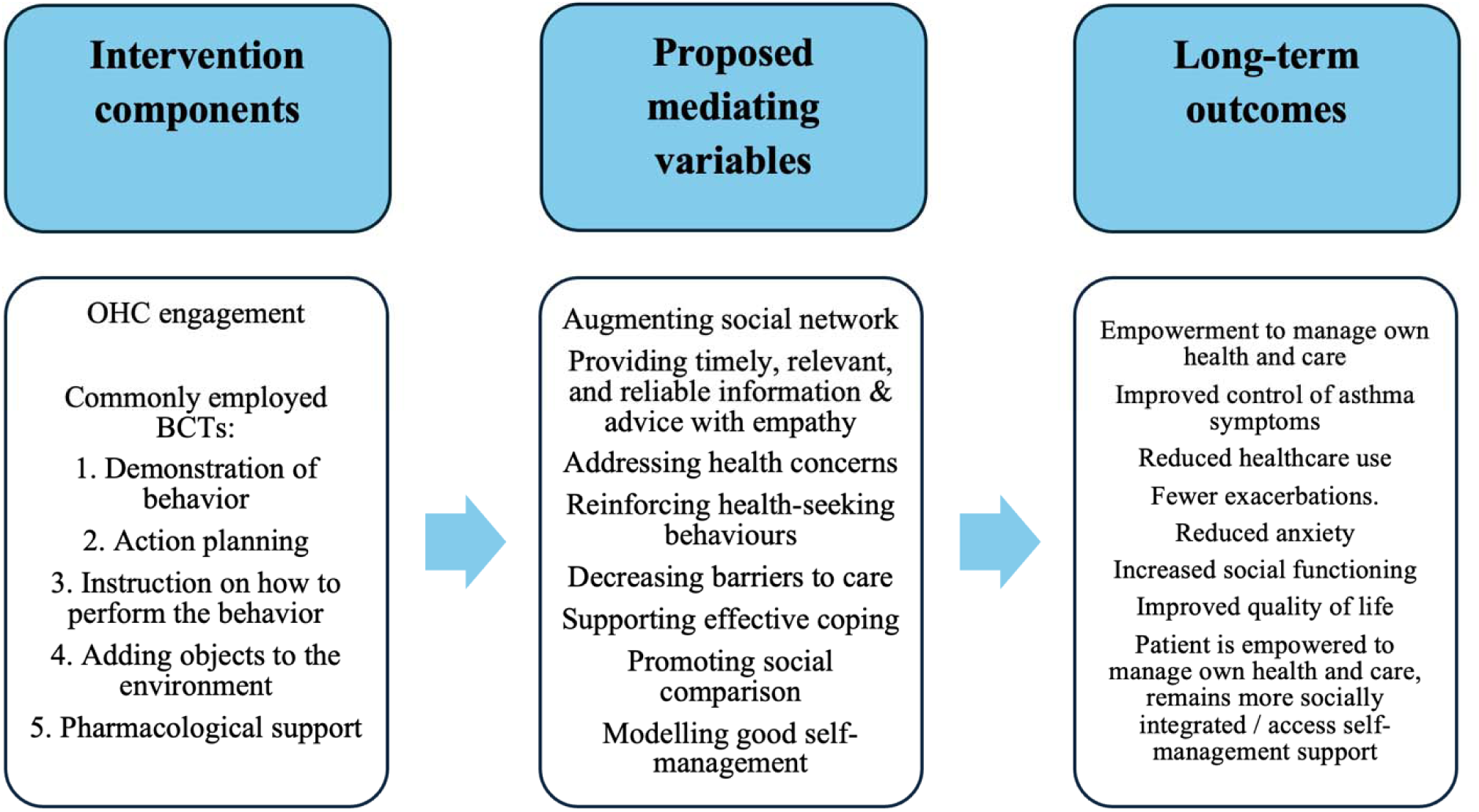
Logic model

There is emerging evidence of how digital media can influence behavior change. Previous studies show that OHCs can have a positive impact on health-related behavioral change [8,14]. Few studies have explored determinants of behavior change in OHCs. One study has shown that social integration, descriptive norms and social support from online social relationships have a positive association with users’ health behavior [14]. Other studies have shown that OHCs have a positive impact on promoting healthy behaviors such as exercise and reducing smoking [18,19]. The current study demonstrates that peer support via an OHC can promote self-management behavior among asthma patients and potentially reduce healthcare use by helping patients to identify situations in which professional healthcare advice should be sought, versus when self-management behavior alone is appropriate. Our findings also shed light on the mechanisms via which online peer support can influence behavior change, by using specific BCTs.

### Implications for healthcare professionals (HCPs) and future research

Raising HCPs’ awareness of OHCs as a potential source of information, advice and support can further promote self-awareness and improve self-management of people with chronic conditions such as asthma [30]. Once proven to be cost-effective to improve health outcomes through a robust randomised controlled trial, healthcare systems could integrate OHCs such as ALUK into their routine care by signposting diagnosed patients to them as a source of information and support. Our team has submitted a trial protocol to co-design and evaluate an intervention to promote engagement with this asthma OHC [29].

The findings of this study can provide valuable insights for OHC moderation teams, helping them to guide and facilitate behavior change in participants to improve self-management. This may include formal training in the use of BCTs, and how a combination of BCTs in their replies rather than using a single one can help prompt behavior change in users. This would allow the moderation team to tailor their responses to encourage behavioral change in accordance with the OHC guidelines. The findings also provide insight into common types of advice that forum users require, for example contacting a clinician or ALUK helpline. This can help improve how resources/signposting given to the users, for example, by creating pinned posts with resources, or external weblinks.

Further research is needed to provide OHC moderators with guidance on how to effectively respond to users’ posts and optimize their advice to maximize encouragement of behavior change. For example, moderators could recommend that users adopt BCTs most likely to facilitate change, such as action planning.

Further research across other OHCs is also needed to determine whether our findings are applicable and can inform self-management strategies for individuals with other chronic conditions.

Associations between positive/negative sentiment and behavior change in OHCs might also be worth exploring, for example, by using semi-automated AI-driven approaches like sentiment analysis. This information could offer valuable insights for community administrators and HCPs, such as nurses, to enhance their support and guidance for users.

## Conclusion

Patients with chronic illnesses are increasingly engaging with OHCs for support and advice. We found qualitative evidence of instances of self-management behavior change and intention to change behavior resulting from engagement with an asthma OHC, with Behavior Change Techniques being widely employed by users offering advice. We identified ‘action planning’, ‘instruction on how to perform the behavior’, ‘adding objects to the environment’, and ‘pharmacological support’ as effective techniques influencing the self-management behavior of asthma patients. Our findings shall give confidence to healthcare professionals about promoting online peer support to well moderated asthma OHCs, e.g., during discussions with their patients about obtaining health information online. As such, these findings are likely to enhance participation in and use of OHCs, thereby enhancing the potential of OHCs in terms of fostering self-management behavioral change.

## Acknowledgements

We are grateful to Asthma UK and HealthUnlocked for granting permission to analyze the online community.

VD was partly funded through Research Capability Funding. This study was partly funded by a Programme Grants for Applied Research Programme (grant number: NIHR202037). Views articulated are those of the authors and not necessarily those of the National Health Service, NIHR, or the Department of Health and Social Care.

The authors attest that there was no use of generative artificial intelligence (AI) technology in the generation of text, figures, or other informational content of this manuscript.

## Data availability statement

The datasets generated or analyzed during this study are not publicly available as they can easily be used to identify the participants, but are available from the corresponding author upon reasonable request.

## Authors’ contributions

Dhir V - Investigation, writing - original draft

Wood HE - Investigation, validation, writing - review & editing

Li X - Formal analysis

Karampatakis GD - Methodology, validation

Griffiths CJ - Conceptualization

De Simoni A - Conceptualization, Methodology, Funding acquisition, Project administration, Supervision, writing - review & editing

## Ethical Approval

Ethics approval for this study was assessed by the Queen Mary’s University Research Ethics Committee and was exempt from full review (QMERC2020.060).

## Abbreviations

ALUK: Asthma + Lung UK
BCT: Behaviour change technique
BTS: British thoracic society
HCP: Health care professional
NICE: National institute for health and care excellence
OHC: Online health community
OTC: Over the counter
SIGN: Scottish intercollegiate guidelines network

